# Patient involvement in the biopsychosocial model of care integrated with primary health services: Experience from three health districts in South Kivu, Democratic Republic of Congo

**DOI:** 10.1101/2024.09.05.24313096

**Authors:** Bertin Mutabesha Kasongo, Christian Eboma Ndjangulu Molima, Gérard Jacques Mparanyi, Samuel Lwamushi Makali, Pacifique Lyabayungu Mwene-Batu, Albert Mwembo Tambwe, Hermès Karemere, Ghislain Balaluka Bisimwa, Abdon Mukalay wa Mukalay

**Affiliations:** Ecole Régionale de Santé Publique (ERSP), Catholic University of Bukavu, Bukavu, Democratic Republic of Congo; School of Public Health (ESP), University of Lubumbashi, Lubumbashi, Democratic Republic of Congo; Faculty of Pharmaceutical Sciences and Public Health, Official University of Bukavu, Bukavu, Democratic Republic of Congo; Centre de recherche sur les politiques et systèmes de santé (CR3-POLISSI), Ecole de santé publique, Université libre de Bruxelles, Brussels, Belgium; Centre de Recherche en Sciences Naturelles, Lwiro, Democratic Republic of Congo; Université du Cinquantenaire, Lwiro, Democratic Republic of Congo

**Keywords:** patient, involvement, health policy, biopsychosocial model, health district, DRC

## Abstract

**Introduction:** Involving people in the provision of care and in decision-making about their health is one of the keys to success and to improving service delivery in the provision of quality health care. Patient involvement in the biopsychosocial model of care is poorly documented in the Democratic Republic of Congo (DRC). The aim of this study is to describe patients’ involvement in the choice of their health policy, their responsibility in holistic care and their capacity to support the biopsychosocial model.

**Methods:** This qualitative research was conducted in three health districts in the province of South Kivu, DRC. Using a tool inspired by the International Alliance of Patients’ Organizations’ Declaration of Patient-Centered Healthcare, we conducted 27 individual interviews between February and April 2024. These interviews concerned people in complex situations, attending health centers and belonging to patient clubs in six health areas covered by the study. A content analysis of the discourse from the various interviews was carried out.

**Results:** Patient involvement in the biopsychosocial model of care depended on multiple factors, including relational aspects (partnership between providers and patients, discussion of therapeutic possibilities and guidance for choice), educational aspects (advice and teaching received from caregivers, development of skills), empowerment (responsibility for care), organizational aspects (inclusive and participatory planning, access to different health services) and community aspects (role of entourage and patient clubs).

**Conclusions:** The various factors influencing patient involvement in the BPS model should be taken into account in the definition of policies and the process of integrating biopsychosocial care. Some of the strategies suggested to support the model, such as raising awareness of the humanization of care, improving the availability of resources and strengthening financial autonomy, will help to improve the quality of care and accessibility to quality health services.

## Introduction

Involving people in the provision of care and in decision-making about their health is one of the keys to success and to improving service delivery in the provision of quality health care. (1,2). Involving people in their own care means allowing them to participate autonomously in defining the health care policies that concern them, taking into account their needs, preferences and independence (3–5).

Various strategies and models for explaining people’s involvement in care have been provided worldwide. These include health policy development, participation in decision-making about their health, co-production of health services, quality improvement and restructuring of health systems… (6–8).

Some studies have revealed that patients who are involved in the provision of care express a sense of well-being and self-satisfaction (9). Others have reported improved interaction and communication between providers and patients (10), patient adherence to treatment (11), and consideration of patients as partners in care(12).

In the context of low-income countries, particularly in Sub-Saharan Africa, the participation and involvement of patients in decision-making about their care is crucial. The aim is to improve both their health and the health systems, which are still fragile. In some situations, this involvement is limited to service design and health research (13), or patients’ ignorance of their right to participate in the decision-making process in their care, especially the less educated (14). Other experiences are either conclusive in terms of positive results in the provision of person-centered care (PCC), as in the treatment of HIV (15). Some provide strategies for involving patients in their own care and thus facilitating the achievement of these positive outcomes, particularly in mental health (16), where the biopsychosocial (BPS) model of care would also require taking spiritual aspects into account (17). Involving patients in their care also requires empowerment, as shown by these studies analyzing the barriers and facilitators to patient empowerment in controlling their disease (18,19).

In the Democratic Republic of Congo (DRC), particularly in the Eastern region where the health system is faced with complex situations and crises, involving patients in their biopsychosocial care is crucial to the system in order to strengthen collaboration between providers and patients, offering them care adapted to their needs, according to biological, psychological and socio-cultural dimensions, and thus improving the quality of health care.

Various studies have been carried out into patient involvement in decision-making about their health and care services for certain health situations (20,21), their perceptions and also their satisfaction with the holistic care on offer (22,23). However, to our knowledge, patient involvement in the BPS model of care is poorly documented in the DRC.

The aim of this study is to describe patients’ involvement in choosing their health policy, their responsibility for holistic care and their ability to support this BPS model.

## Methods

### Study settings

#### Organizational framework of the health system in the DRC (operational level)

The present study was carried out in South Kivu Province, DRC, more specifically in three Health Zones (HZ) or Health Districts (HD).

The DRC’s healthcare system is divided into three levels: the central or national level, the intermediate or provincial level, and the peripheral or operational level. Our study takes place at the operational level, made up of HDs.

This level is responsible for implementing Primary Health Care (PHC) strategies. It comprises the HDs (mostly with a General Reference Hospital each), which are subdivided into Health Areas (HA), each with a Health Center (HC). The health districts operate under the coordination of the Health Zone Management Team. This coordination involves the planning, implementation, monitoring and evaluation of all HD activities, operationalized at HA level, by the HC team (HC Head Nurse and other care providers). The Health Center is the point of contact between the health system and the population, through community participation structures, direct community representatives (Health Area Development Committee or CODESA, which is one of the HC’s co-management structures, Community Animation Cell – CAC; Community Health Workers – CHWs) and other structures known as “community-based organizations” (organisations à assises communautaires - OAC) (24,25).

#### Geographical and health context of the HDs

The three HDs concerned by our study have been receiving support from Louvain Coopération as part of the Non-Communicable Diseases Program (PMNT) since 2022. This project focuses on the management of diabetes, hypertension and mental health, as a gateway to BPS care. Table 1 gives some health information for these three HDs.

**Table 1.** Description of health districts.

| Health District | Population<br>(inhabitants) | HA/HC<br>Number | Hospital | Rates of utilization of curative<br>services (%)* |
| --- | --- | --- | --- | --- |
| Bagira | 162186 | 8 | 3 | 32.8 |
| Katana | 267504 | 20 | 4 | 72.1 |
| Walungu | 318610 | 23 | 3 | 32.5 |

Source: health pyramid of South Kivu HD and DHIS2, 2023
\* The health standard for the rate of utilization of curative services is $\geq 50\%$

This program covers four HA in each of these three HD. For the purposes of this study, six of the twelve HAs benefiting from this support were selected on the basis of reasoned choice and accessibility, with 2 HAs per HD (Lumu and Nyamuhinga HAs in the Bagira HD, Birava and Kabushwa HAs in the Katana HD, and Bideka and Lurhala HAs in the Walungu HD).

#### Study design and period

This was a cross-sectional study, using a descriptive qualitative approach (26,27), carried out during the period February to April 2024.

#### Conceptual framework for community engagement in the BPS model of care

This study is based on the conceptual framework of community involvement in the biopsychosocial model of care, which we have developed. Community involvement in care according to the BPS model requires interaction between the community and the healthcare system, represented by the provision of integrated services. We have developed the various components to be taken into account for the biopsychosocial perspective (availability of qualified health personnel, quality health services, social groups and the individual).

This framework is represented in Fig 1.

**Fig 1:**
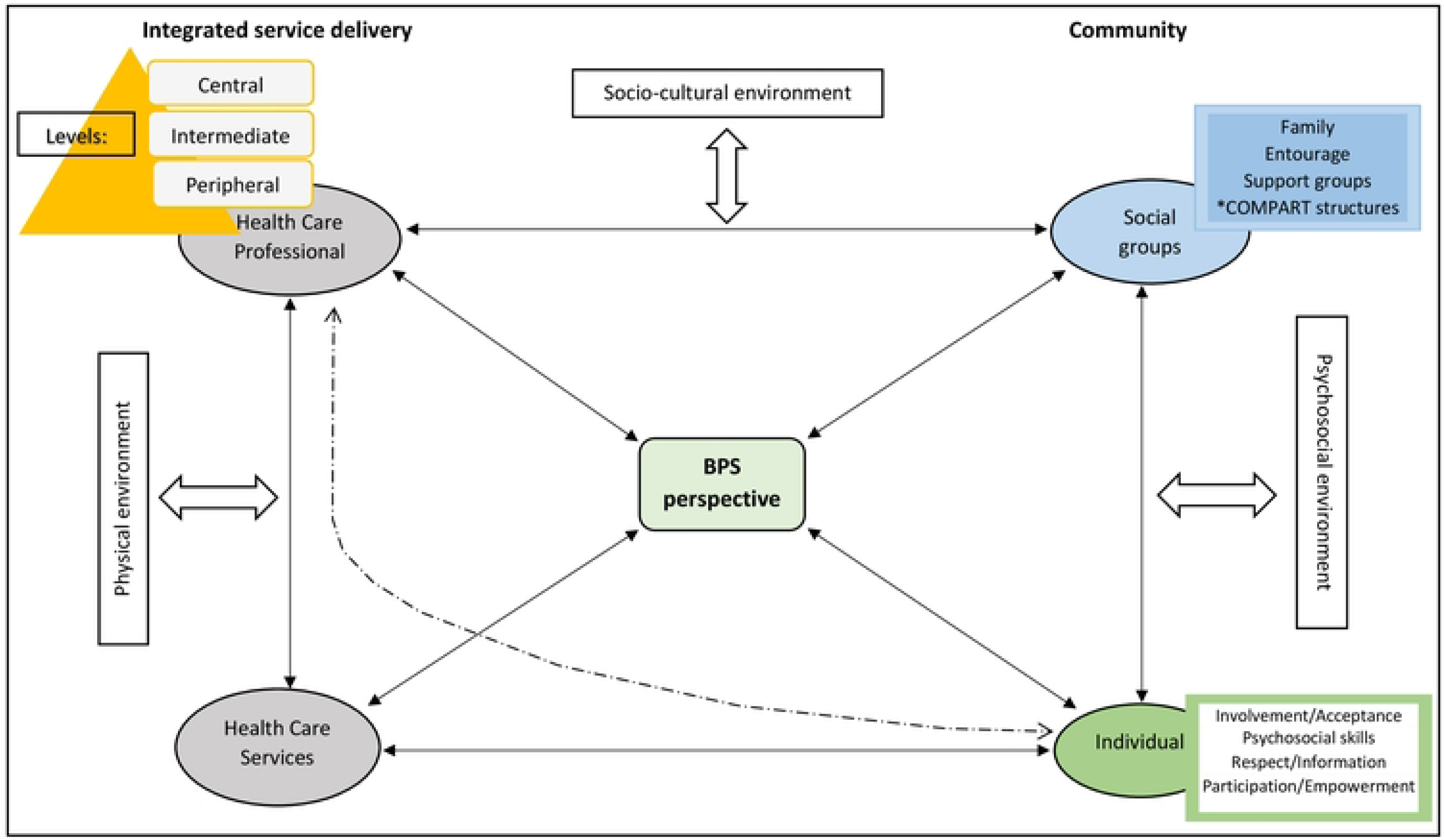
Conceptual Framework for Community Engagement in the BPS Model (*COMPART : Community Participation)

In this sense, on the health system side, it will be necessary to act on the physical environment, in particular:

- The availability of qualified healthcare staff: on the front line of care, not only are the acts performed by staff complementary, but multidisciplinarity should also be encouraged.
- Availability of quality healthcare services: quality care requires both qualified personnel and good quality healthcare services.

For the community, we need to act on the psychosocial and sociocultural environment, taking into account the individual and social or community groups:

- Individual: the person who is responsible for his or her own health. In this context, “empowerment” is defined as described in health promotion (28,29). A key element of the BPS model is the individual’s participation in the provision of care. His recovery depends on his involvement or acceptance in the provision of care (2). Elements directly related to the individual are also taken into account, including biological and genetic characteristics, personal and social skills, lifestyle habits and behaviors, and socioeconomic characteristics.
- Social groups: here we have all the community groups and social networks, starting with the person’s family and entourage, which are the primary social groups, with a major socio-cultural influence on health (30,31). Then there are the various associations and groups, including the diabetics’ club, the club for people living with AIDS (considered to be community-based organizations - OAC), as well as community participation bodies such as CODESA, CACs…, all of which provide immediate support to the individual. These elements need to be taken into account in care, especially in psychosocial support, a key component of the BPS model of care (32,33).

In this study, we focus on the “individual” component (in green in Figure 1) and its interactions with the other components, in particular its involvement in the definition of policies related to decision-making in BPS care, the integrated service delivery component having been the subject of other studies elsewhere (34,35). The “social groups” component has been developed in another complementary work.

The conceptual model presented above is inspired by the theoretical frameworks of person-centered care proposed by De Man J et al.(36), Moreau A et al. (37) and Mead N & Bower P (38).

#### Study participant selection

A selection by convenience was made to include participants in the study. We took into account patients who belonged to clubs (diabetic and hypertensive clubs), considered to be patient support groups. We considered patients who had been members of these clubs for at least one year prior to the survey, and who were active in the activities organized by these clubs. This was done in order to understand the dynamism around the BPS model and the interaction between the three components of our conceptual framework, namely the individual, the healthcare professional and social groups (specifically support groups). A total of 27 patients were selected for this study.

Table 1 shows their distribution in different HDs.

**Table 1:** Persons interviewed on the basis of origin.

| Health District | Health Area | Persons interviewed |
| --- | --- | --- |
| Bagira | Lumu | 4 |
|  | Nyamuhinga | 5 |
| Katana | Birava | 5 |
|  | Kabushwa | 5 |
| Walungu | Bideka | 5 |
|  | Lurhala | 3 |
| Total |  | 27 |

#### Data collection

We conducted individual semi-structured interviews with selected patients. Some were met at the HC when they came for treatment, others were found in their households.

An interview guide based on the International Alliance of Patients’ Organizations’ Declaration of Patient-Centered Care (39) was used to collect data.

The interviews focused mainly on describing the relationship between patients and providers on the one hand, and between patients and the community on the other (mainly support groups); the respect for patients’ needs, autonomy and independence by providers in offering care; patients’ involvement in the choice of care policies in relation to their health and in decision-making; their share of responsibility in holistic care; their ability to support the approach; where possible, to propose solutions for maintaining this BPS care.

These interviews were conducted by two members of the research team, after obtaining the interviewees’ consent and explaining the purpose of the study.

They were conducted in Swahili and/or the local language (Shi), with an interpreter facilitating translation, and lasted an average of 45 minutes. It should be noted that there was only one interpreter for all these interviews, who spoke, understood and wrote the local language well. The interviews were recorded on an audio recorder, then transferred to a laptop for storage.

#### Data analysis

The recorded interviews were transcribed verbatim into Word files before being translated into French, using two independent translators to ensure the accuracy of the interviewees’ words.

We then carried out a content analysis of these transcripts, using a combination of inductive and deductive approaches (40). We first familiarized ourselves with the interviews, which we read and reread, before identifying and coding the content that had been discussed and agreed upon. We then grouped them according to repetition and similarity, and associated them with categories deduced globally from the principles of the International Alliance of Patients’ Organizations’ Declaration of Patient-Centered Care, which we considered as sub-themes (41). Finally, we defined four main themes, by consensus, by grouping together the sub-themes retained, after team discussions: (1) Participation and Empowerment; (2) Involvement and Acceptance; (3) Psychosocial Skills and (4) Respect and Information. The following table (Table 2) provides an illustration of the data analysis process. Our data are presented according to these selected sub-themes and main themes, supported by a few verbatims. BKM, GJM and CENM participated in this data analysis. We have taken into account in this analysis the categories identified that were contradictory to those mainly retained.

**Table 2:**
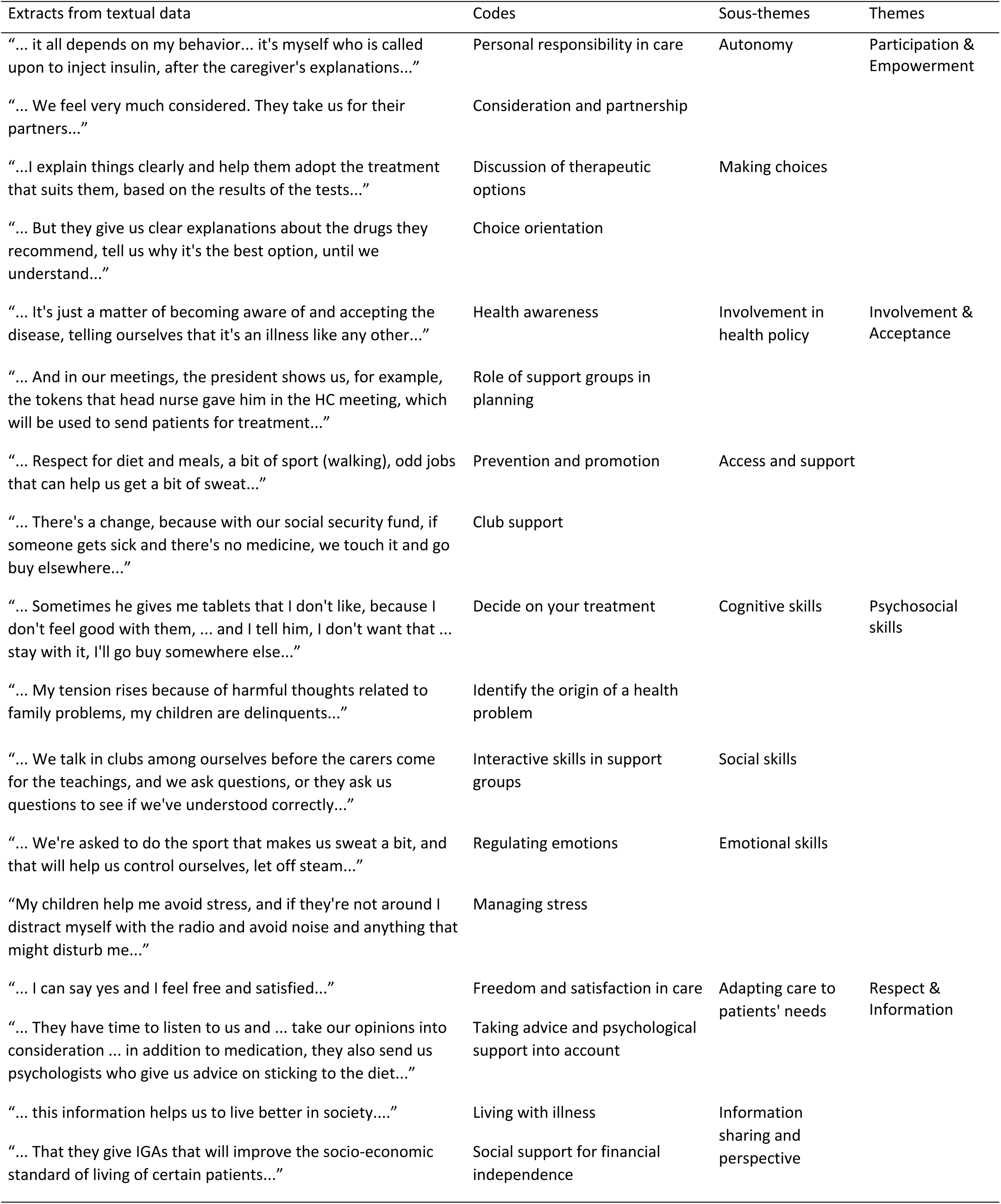
Illustration of the data analysis process.

| Extracts from textual data | Codes | Sous-themes | Themes |
| --- | --- | --- | --- |
| "... it all depends on my behavior... it's myself who is called upon to inject insulin, after the caregiver's explanations..." | Personal responsibility in care | Autonomy | Participation & Empowerment |
| "... We feel very much considered. They take us for their partners..." | Consideration and partnership |  |  |
| "...I explain things clearly and help them adopt the treatment that suits them, based on the results of the tests..." | Discussion of therapeutic options | Making choices |  |
| "... But they give us clear explanations about the drugs they recommend, tell us why it's the best option, until we understand..." | Choice orientation |  |  |
| "... It's just a matter of becoming aware of and accepting the disease, telling ourselves that it's an illness like any other..." | Health awareness | Involvement in health policy | Involvement & Acceptance |
| "... And in our meetings, the president shows us, for example, the tokens that head nurse gave him in the HC meeting, which will be used to send patients for treatment..." | Role of support groups in planning |  |  |
| "... Respect for diet and meals, a bit of sport (walking), odd jobs that can help us get a bit of sweat..." | Prevention and promotion | Access and support |  |
| "... There's a change, because with our social security fund, if someone gets sick and there's no medicine, we touch it and go buy elsewhere..." | Club support |  |  |
| "... Sometimes he gives me tablets that I don't like, because I don't feel good with them, ... and I tell him, I don't want that ... stay with it, I'll go buy somewhere else..." | Decide on your treatment | Cognitive skills | Psychosocial skills |
| "... My tension rises because of harmful thoughts related to family problems, my children are delinquents..." | Identify the origin of a health problem |  |  |
| "... We talk in clubs among ourselves before the carers come for the teachings, and we ask questions, or they ask us questions to see if we've understood correctly..." | Interactive skills in support groups | Social skills |  |
| "... We're asked to do the sport that makes us sweat a bit, and that will help us control ourselves, let off steam..." | Regulating emotions | Emotional skills |  |
| "My children help me avoid stress, and if they're not around I distract myself with the radio and avoid noise and anything that might disturb me..." | Managing stress |  |  |
| "... I can say yes and I feel free and satisfied..." | Freedom and satisfaction in care | Adapting care to patients' needs | Respect & Information |
| "... They have time to listen to us and ... take our opinions into consideration ... in addition to medication, they also send us psychologists who give us advice on sticking to the diet..." | Taking advice and psychological support into account |  |  |
| "... this information helps us to live better in society..." | Living with illness | Information sharing and perspective |  |
| "... That they give IGAs that will improve the socio-economic standard of living of certain patients..." | Social support for financial independence |  |  |

#### Ethics statement

This research was approved by the Ethics Committee of the Catholic University of Bukavu (UCB) under the order number UCB/CIES/NC/002/2024. The study took care to comply with the following ethical principles, in line with the WMA Declaration of Helsinki. Prior to data collection, the researcher was careful to explain the reasons and objectives of the study. He then presented the interviewees with an informed consent form, which they were invited to read and sign before the interview began. All respondents voluntarily accepted and signed the written consent form, consented to participate in the study and the audio recording. Respondents were free to express themselves in any language they felt comfortable with (Swahili or Shi), with an interpreter to facilitate translation for interviews in the local language (Shi). Respondents were assured anonymity in order to respect their confidentiality.

## Results

### 1. General characteristics of participants

Table 3 shows the general characteristics of the study participants. The majority of respondents were female (67%), almost half were aged between 45 and 65. 67% had attended at least elementary school and most (41%) were without formal employment (unemployed and housewives).

**Table 3:**
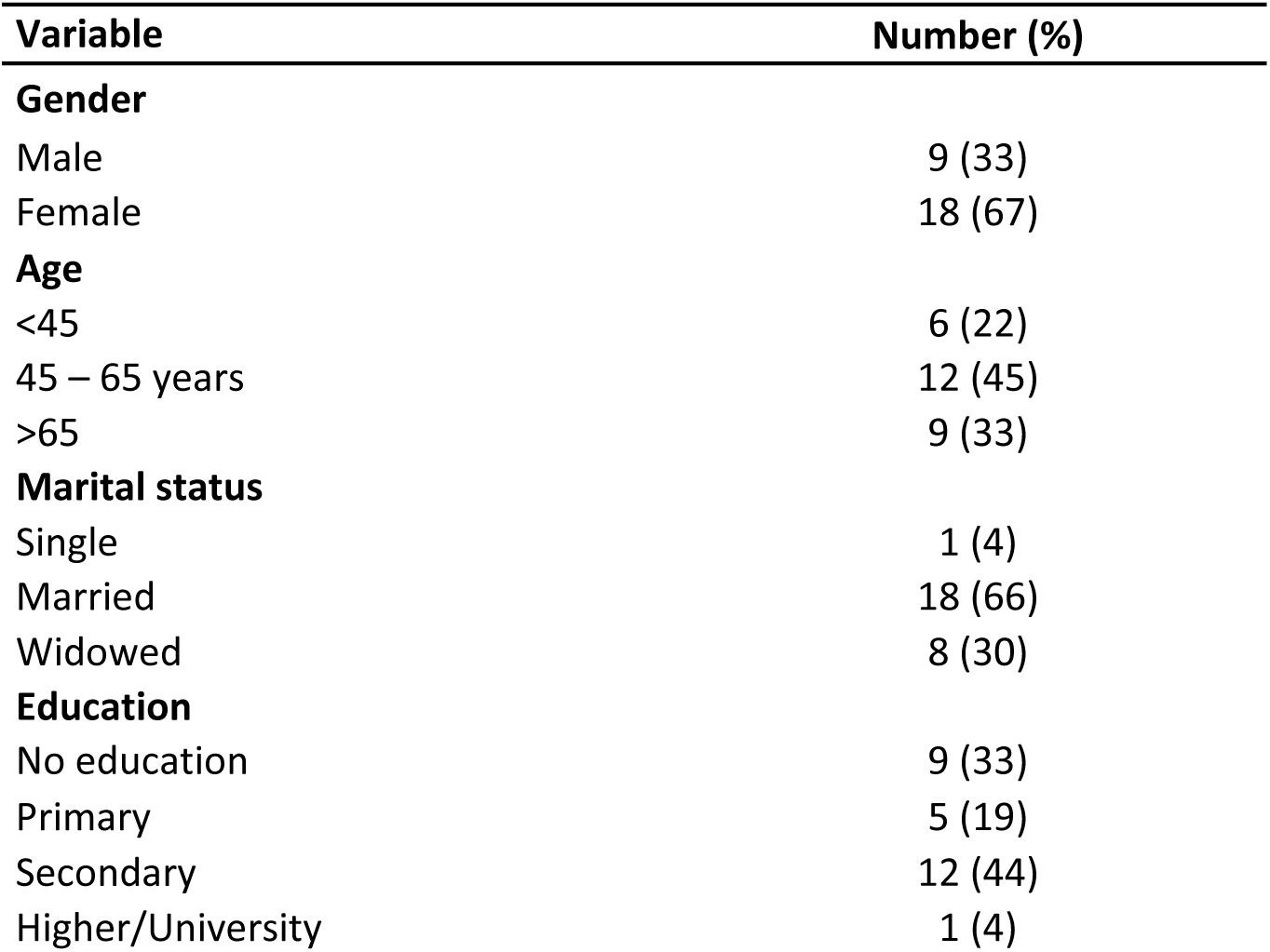
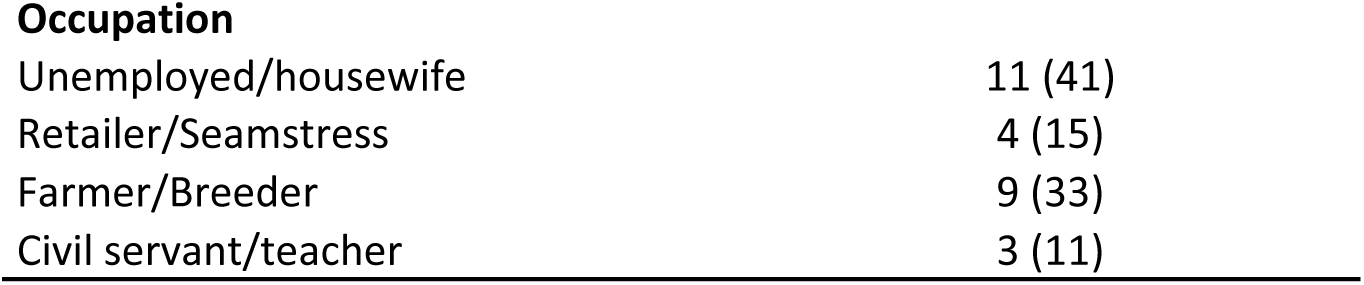
Respondents’ general characteristics.

| Variable | Number (%) |
| --- | --- |
| **Gender** |  |
| Male | 9 (33) |
| Female | 18 (67) |
| **Age** |  |
| <45 | 6 (22) |
| 45 – 65 years | 12 (45) |
| >65 | 9 (33) |
| **Marital status** |  |
| Single | 1 (4) |
| Married | 18 (66) |
| Widowed | 8 (30) |
| **Education** |  |
| No education | 9 (33) |
| Primary | 5 (19) |
| Secondary | 12 (44) |
| Higher/University | 1 (4) |
| Occupation |  |
| Unemployed/housewife | 11 (41) |
| Retailer/Seamstress | 4 (15) |
| Farmer/Breeder | 9 (33) |
| Civil servant/teacher | 3 (11) |

### 2. Participation & Empowerment in the BPS model

#### 2.1. Autonomy

Patients recognize their responsibility in maintaining their health, by following the instructions of their caregivers, particularly when it comes to self-managing treatments such as insulin.

> *“… everything depends on my behavior, I’m called upon to follow instructions, instructions: for example, it’s myself who’s called upon to inject myself with insulin, after the caregiver’s explanations.” (Patient Bideka 5)*

Some also say that they feel considered, and see that there is good interaction between them and the providers, who see them as partners. This motivates them and reassures them about the difficulties they personally face. *“We talk to the nurses, we tell them about our worries, our problems and everything we’d like… they also take them into consideration and help us…”. (Patient Lumu 5)*

Others, on the other hand, find that this consideration is not noticeable among certain providers who display a certain negligence towards them, which frustrates patients.

> *“Only one thing that bothers us is the lack of the Head Nurse involvement in our activities like other caregivers do. We’re even beginning to think that he’s discrediting us” (Patient Kabushwa 4).*

#### 2.2. Making choice

In their care, patients are actively involved in choosing treatments in collaboration with providers. They discuss treatment options, and caregivers help them understand the best options based on test results and their individual needs, as explained by one interviewee, who is a caregiver and part of the club:

> *“… Besides, I’m in charge of it as a caregiver, while being part of the patients. We discuss how to deal with each case, and I guide them… we start with the teachings and then various examinations. I explain things to them clearly and help them to adopt the treatment that suits them, and it’s according to the results of the examinations that we give them medication.” (Patient Kabushwa 1)*

> *“Your state of health itself will define the treatment to be followed (insulin or tablets). But they give us clear explanations about the medicines they recommend, tell us why it’s the best option, until we understand.” (Patient Birava 5)*

## 3. Involvement & acceptance

### 3.1. Involvement in health policy

Most of those surveyed had a positive perception of the relationship between patients and care providers at the facility level. Patients thus participate autonomously in decision-making about the care that affects their lives, taking into account the treatment options available.

> *“They don’t need to impose anything on me because I know that situations differ, that sometimes I have to be hospitalized and take insulin and other times it’s just tablets; however, the nurse can suggest insulin but with the effects it has on us (asthenia and intense hunger), I suggest that they give me tablets and the advice that follows… They accept and tell me anyway that the best thing was insulin… “. (Patient Bideka 1)*

From the advice and teachings receive from healthcare providers, they come to be aware of and accept their health condition, and this enables them to be psychologically stable.

> *“It’s just a matter of becoming aware of and accepting the illness, telling yourself that it’s an illness like any other and respecting the instructions, advice of the caregiver.” (Patient Lumu 1)*

Family and entourage, including support groups, play a crucial role in offering encouragement and advice, thus reinforcing BPS care: *“My wife accompanies me in my illness and always encourages me to go to the health center, even if it’s just a simple malaria. She’s too involved… We also talk to my family about my state of health, and even with some of my neighbors; and they also give me advice, if possible” (Patient Bideka 1)*.

The extent to which clubs are involved in decision-making varies from place to place. In some HAs, respondents report that the leaders of their support groups (the clubs) are involved in planning at the HC level, where the various health activities are discussed with the different stakeholders. At their club meetings, they discuss the activities that concern them and whose implementation they must monitor.

> “There’s the president, and those who come after him who discuss with Head Nurse about our care. And in our meetings, the president shows us, for example, the tokens that the Head Nurse gave him in the HC meeting, which will be used to send patients for treatment” (Patient Bideka 3).

In collaboration with providers, CHWs and club presidents make home visits (VADs) to patients’ households to monitor care, preventive and promotional activities as recommended by providers. Some club members are also CHWs, which further motivates their involvement, and improves interaction between them and the providers.

> “I myself am a CHW. We organize VADs with the Head Nurse, and often I go with our club president to see other members in their households, and there we talk together about their health. We are also informed by the nurse who is in charge of accompanying us, and we participate in HC activities. For example, if there’s a visitor, vaccination or construction activities…”. (Patient Birava 1)

On the other hand, other respondents expressed a lack of involvement in HC activities, apart from awareness-raising sessions by the psychosocial agent during their club meetings.

> “We’re with the Psychosocial Agent (PSA) every time. She teaches us a lot, and participates in our activities. We meet with her on the 24th of every month. Apart from that, they do a lot of things without involving us…” (Patient Kabushwa 3)

### 3.2. Access to health services and patient support in BPS care

Patients benefit from comprehensive treatments that include, in addition to medication, psychological counseling, dietary recommendations and adapted physical activities.

> “Respect for diet and meals, a bit of sport (walking), small jobs that can help us get a bit of sweat…” (Patient Lurhala 3)

During consultations at health care facilities, providers and patients work together to establish a link between the current state of health and other factors such as education, employment, family problems, etc. *“. Sometimes problems arise in the family and we think, we say to ourselves such and such a child is acting badly, or this and that, and other times we’ll think about it all night and in the morning the blood pressure figures go up.” (Patient Bideka 3)*

Patients receive a great deal of support from the social groups to which they belong. They find that these groups bring great added value to this BPS care, especially for psychological and social aspects, such as financial access to medication when needed.

> *“… There’s a change, because with our social security fund, if someone falls ill and there’s no medicine, we touch it and go and buy elsewhere. This club also helps us to be close to each other. At first you may think you’re the only one who’s ill, but when you meet the others, it helps you psychologically; personally, I recently lost my father, and the group really helped me. We also give advice on diet, behavior…”. (Patient Kabushwa 3)*

## 4. Developing psychosocial skills

### 4.1. Cognitive skills

Patients have acquired important skills, thanks to the education they receive in their clubs, including knowing how to decide on their treatments, according to their state of health. *“Sometimes he gives me tablets I don’t like, because I don’t feel good with them, the little ones, and I tell him I don’t want that. If there isn’t the big one there, I tell him, stay with that, I’ll go and buy somewhere else…” (Patient Nyamuhinga 3)*

They are also able to identify the origin of the disturbance in their health, thanks to the information they receive from providers and animators in the clubs they attend, most of which is linked to family problems: *“My [blood] pressure goes up because of harmful thoughts linked to family problems, my children are delinquents…” (Patient Birava 2)*.

### 4.2. Social skills

Support groups enable patients to develop social skills through regular discussion and exchange with peers and facilitators. At their club meetings, they acquire the ability to interact and discuss their health together.

> *“… we talk to each other in clubs before the caregivers come to teach us, and we ask questions, or they ask us questions to see if we’ve understood correctly… In fact, I’m beginning to miss the others who are ill but hiding in the house. There’s a difference between those who stay at home and us who come here. Those who are at home can’t benefit from these teachings…” (Patient Nyamuhinga 2)*

### 4.3. Emotional skills

Some respondents report their experiences in regulating their emotions or managing the stress they face on a daily basis. *“My children help me to avoid stress, and if they’re not around I distract myself with the radio and avoid noise and anything else that might disturb me…” (Patient Lurhala 3)*

The practice of promotional activities, which are carried out in the community on the recommendation of providers, is also reported, to self-regulate.

> *“We’re asked to do the sport that makes us sweat a bit, and that will help us control ourselves, let off steam. For example, I have a friend in MUGOGO, I wake up in the morning, take the road to MUGOGO and walk back.” (Patient Bideka 3)*

However, efforts need to be made to promote group promotional activities, including group sport or physical exercise, which is one of the weaknesses identified by respondents.

## 5. Respect & Information

### 5.1. Adapting care to individual needs

Patients report that the care they receive respects their needs, preferences and independence. Some even say they feel free and satisfied with the treatment they receive at HC.

They are cared for as a whole, physically, psychologically and socially. This last aspect is more noticeable in the community, in the support groups to which patients belong. In some facilities, the hospital psychologist is sometimes called in for the psychological aspects, in the absence of a PSA.

> *“The providers here treat us well; they don’t impose anything on us. They take the time to listen to us and … take our opinions into consideration; they talk to us well, … in addition to medication, they sometimes send us the psychologists who give us advice…” (Patient Lumu 2)*

### 5.2. Information sharing and perspective

Patients receive the information they need for their treatment, from providers and social groups. They discuss together, both in their groups and with caregivers and CHWs, how to live with their disease. Some even exchange experiences with other clubs, and the information they receive from these exchanges helps them to thrive.

> *“Here recently, our club and Burhiba’s club organized a meeting with an exchange of experience.” (Patient Lumu 2)*

> *“…we discuss it in our club and we exchange experience, and we learn the lesson together and measures that help us in our lives, this information help us to live better in society.” (Patient Kabushwa 2)*

To maintain this BPS care, the respondents gave some proposals for its improvement:

- Raising awareness among providers who show indifference
- Building capacity for better care
- Improving the supply of medicines to avoid shortages.
- Increased availability of psychologists for better care, especially for the psychological aspect.
- The strengthening of social funds to support income-generating activities, contributing to greater financial autonomy for patients. *“IGAs (Income-Generating Activities) should be provided, which will improve the socio-economic standard of living of some patients, because treatment also involves diet, but some are unable to adhere to it, or don’t even eat, and may even lose their lives. (Patient Kabushwa 5)*

## Discussion

The results of our study show the importance of patient involvement in their care, their ability to make informed decisions, and the key role of support groups in the success of biopsychosocial management.

The study provided us with various factors influencing this patient involvement in this model, including the partnership relationship between providers and patients, exchanges on therapeutic possibilities and guidance for choice, teaching received from caregivers, skills development, patient empowerment in care, organizational aspects (inclusive and participatory planning, access to different health services) and community aspects (role of entourage and patient clubs). Patients proposed strategies to support the model, including the humanization of care, capacity-building for providers, psychological assistance and the implementation of activities facilitating financial autonomy.

### The patient-caregiver relationship and empowerment in integrated and comprehensive care

Positive interaction between health service providers and users generates a feeling of consideration and motivation to become more involved in care, as the results of our study show. They also declare that the care offered to them is comprehensive, tailored to their needs, desires and independence, and testify to a sense of satisfaction with the treatment they receive. An analysis of the expected and perceived satisfaction of users of VVS (victims of sexual violence) services for holistic care at Panzi Hospital, eastern DRC, identified various factors influencing satisfaction, depending on the organizational framework of the services provided as well as the social environment of the beneficiaries (22).

Others, on the other hand, have found that some providers are indifferent to their gaze, without consideration, and display a certain neglect towards them, leading to frustration on their part. Lack of empathy and compassion towards the patient can have psychological repercussions, leading to feelings of dissatisfaction, rejection and lack of confidence, all of which have a negative impact on therapeutic success. This should be taken into account in the biopsychosocial model of care (42,43).

The empowerment of care is an essential factor to consider in the BPS model of care. We have seen from our respondents that the patient has an important role to play in BS care, as the person responsible for his or her own health. He or she is simply called upon to follow the instructions and advice provided by providers to maintain his or her health.

Other elements that reinforce this provider-recipient relationship and empowerment are the discussions that take place either during the consultation or during the educational sessions, in relation to the various therapeutic possibilities and various other advice on how to live with their illness, as mentioned by our respondents. They also receive guidance in choosing the option that seems best, taking into account their needs. These educational sessions also enable them to develop their knowledge and acquire skills. In this context, we find the elements of health literacy, notably functional knowledge (how to inject insulin, for example), interactive knowledge (the discussions they have in their meetings) and critical knowledge (discussions on the different therapeutic possibilities and making the choice), as well as elements of psychosocial skills development (the ability to identify, for example, the origin of the disturbance in their state of health, experience in regulating their emotions or stress management through preventive and promotional activities, including sport…). Other authors have tried to analyze these elements of literacy and psychosocial skills in the patient care process (44–46).

We also felt that efforts needed to be made to strengthen group promotional activities, including group physical exercise, which was one of the weaknesses identified by our respondents. These types of exercise can be carried out under the supervision of providers. The experience of the Centres de Santé Médicalisés Urbains (CSMU) in the east of the DRC is an example of successful care and user satisfaction, thanks to the availability of a physiotherapy service to promote these physical activities under medical supervision (47).

### Participation/Engagement of patients in health policy as partners in BPS care

Partnership in care means involving patients in the definition of policies concerning their health, involving them in planning and decision-making, in order to reassure themselves that their needs are being taken into account, that they are being treated with dignity, and thus contribute to improving the quality of care. Most of the people we interviewed said that providers see them as partners, which reassures them of the difficulties they face personally.

This partnership is also felt by the entourage and support groups (patient clubs) who are involved in biopsychosocial care, through the accompaniment and discussions they have with caregivers and the follow-up in care. Club leaders are also involved in planning the various health-related activities at HC level with the various other stakeholders, and discuss with their members the activities that concern them and whose implementation they must monitor.

This partnership is one of the key factors in user satisfaction, as revealed by Heberer M et al. (48). Pomey M-P et al. show us in their study on the contribution of patients to improving the quality of healthcare services, the advantage of involving patients in decision-making, the care process and the implementation of improvement actions, by drawing on their own experience (10). Other countries have set up patient partner organizations to help improve care and achieve better health outcomes (49). Abelson J et al., through a survey organized in Canada, in trying to understand this partnership, were able to identify factors that facilitate or hinder it, and gave guidelines for the success of this collaboration (50).

### Access to information and other care services and support for the BPS model

In terms of access to information, we noted among our respondents that patients receive it in relation to their treatment, within the clubs to which they belong (discussion sessions with facilitators and caregivers). In these patient clubs, they find the time to share their experiences, give each other advice, and the providers also come to hold educational themes on the daily management of the state of health, hygiene, adherence to diet, preventive and promotional measures, how to live with one’s illness… Patients receive a great deal of support from the groups or social networks to which they belong. They find that these groups add a great deal of value to their BPS management, especially in terms of the psychological and social aspects (notably moral support, improved interaction between patients and providers, mutual social assistance…). And all this helps them to flourish in society. These benefits of support systems have also been cited by other authors (33,51,52).

The role of community representatives, including CHWs, is also mentioned by respondents, especially their involvement in VADs, in support of providers, during which exchanges on health status and monitoring of preventive and promotional activities take place. Some authors have demonstrated this role in the management of certain pathologies, notably hypertension and user satisfaction, maternal and neonatal health… (53,54).

However, we did note the desire to see BPS care maintained, through a number of proposals, including raising the awareness and capacity of certain providers, particularly in the area of humanizing care, and strengthening the social fund to implement a number of income-generating activities with a view to financial autonomy. The humanization of care and activities to support financial autonomy have proved their worth in improving health services and the quality of care (55–58).

### Study limits and operational implications

Our study was limited to only one category of patients, presenting complex situations and belonging to the patients’ clubs for at least one year prior to the study. The inclusion of other patients, also those not attending these clubs, would provide additional information, within the framework of this biopsychosocial management. We collected patients’ opinions, without including observation of medical practice with providers, or participation in these support group activities to reassure ourselves of real involvement and interaction in this care process. As noted in the study, the respondents’ level of education could influence the results, especially for the uneducated. The selection criteria mentioned above help to minimize this bias: respondents who have been attending the clubs for at least a year, and who have participated in several educational sessions, appear to be more predisposed to becoming aware of their health status and changing behavior for their well-being.

For respondents who spoke the local language (“Shi”), we used a single interpreter, as this could reduce the comprehensibility or accurate translation of our respondents’ statements, and thus influence the interpretation of the results. Given the size of our sample, the information may not have been diversified. Longitudinal studies, including more people, would enable us to follow the evolution of empowerment and measure this involvement of patients in the BPS model.

The results of our study should encourage health systems to define and adopt policies for involving patients in the provision of care, to promote practices that strengthen the patient-provider partnership, and specifically, in the context of the DRC, a country with limited resources, capitalize on them to improve access to care by strengthening patients’ financial autonomy.

### Conclusions

This study of patients with complex situations in three health zones in South Kivu, DRC, on their involvement in the BPS model of integrated care in primary health services, allowed us to describe this involvement in the choice of their health policy, their responsibility in holistic care and their ability to support this BPS model.

The results of the study identified a number of factors that influence patient involvement in this model, and which should be taken into account in policy-making and the process of integration of biopsychosocial care. Strategies discussed to support the model, including increasing awareness of humanizing care, improving the availability of resources, and increasing financial self-sufficiency, improve the quality of care and access to quality health services. This research should be extended to other health situations, both chronic and acute, in the PCC approach.

## Data Availability

Data cannot be shared publicly because of the confidentiality reassured to respondents. They are available from the institutional data access and ethics committee of the Catholic University of Bukavu for researchers who meet the criteria for access to confidential data.

## Abbreviations

BPS: Biopsychosocial
CHW: Community Health Workers
DRC: Democratic Republic of the Congo
HA: Health Area
HC: Health Center
HD: Health District
IGA: Income-Generating Activity
OAC (Organisation à Assise Communautaire): Community Based Organization
PCC: Person-Centered Care
PSA: Psychosocial Agent;
VAD (Visite à Domicile): Home Visit.

## Acknowledgements

The authors thank all survey respondents for their participation in the study as well as South Kivu Provincial Health Division for facilitating the descent into HDs for data collection.

## Supporting information

S1 Appendix. Interview guide

